# Projected spread of COVID-19’s second wave in South Africa under different levels of lockdown

**DOI:** 10.1101/2021.01.22.21250308

**Authors:** Elisha B. Are, Caroline Colijn

## Abstract

South Africa is currently experiencing a second wave of resurgence in COVID-19 infection. In this modelling study, we use a Bayesian compartmental model to project possible spread of the second wave of COVID-19 in South Africa under various levels of lockdown restrictions. Our model suggests that strict lockdown restrictions will have to be in place up to the end of March 2021 before cases can drop to levels observed, in September to early November 2020, after the first wave. On the one hand, extended lockdown restrictions have negative consequences – albeit effective, they are not sustainable over extended periods. On the other hand, short lockdown restrictions over a few weeks will not have a lasting effect on the spread of the disease. Lockdown restrictions need to be supplemented with increased rapid testing, palliative support for the vulnerable, and implementations of other non-pharmaceutical interventions (NPIs) such as mask mandate. These multifaceted approaches could help keep cases under control until vaccines are widely available.

## 1 Introduction

Globally there have been more than 81,586,011 confirmed cases of infection with novel severe acute respiratory syndrome–coronavirus 2 (SARS-CoV-2 virus), which causes the disease known as coronavirus disease 2019 (COVID-19), with over 1,780,710 reported fatalities to date [1]. In South Africa, as of 28 December 2020, the number of confirmed cases stood at 1,011,871 with more than 27,071 deaths [2]. Owing to recent rises in reported cases, the national department of health (South Africa) officially declared the second wave of COVID-19 in the country, and in response to the resurgence, the government announced adjusted alert level 3 lockdown restrictions, which took effect from midnight of 28 December 2020. The restrictions were initially set to last until 15 January 2021, but restrictions have now been extended infinitely until cases are brought under control. The second wave of COVID-19 will pose a considerable risk to public health and to the socioeconomic well-being of the country.

We aim to project different scenarios for the spread of the second wave of COVID-19 in South Africa using the current levels of contact as a baseline, and to assess possible impacts of various lockdown scenarios on the spread of the disease. Some studies have used mathematical modelling to understand and quantify the spread of COVID-19 in African countries [3,4,5,6,7,8,9], with some of them focusing specifically on the South African context [10,11]. However, as far as we know, this study is the first attempt at predicting the spread of the second wave of COVID-19, and at assessing the impact of lockdown restrictions in South Africa during the second wave.

## 2 Data sources and model description

### 2.1 Data

We use openly available data on reported cases of COVID-19 in South Africa from 5 March 2020 to 27 December 2020. Data are retrieved from the official data released by the National Institute for Communicable Diseases and the Department of Health of South Africa, by the Data Science for Social Impact research group, based at the University of Pretoria, South Africa [12]. The data are available in the GitHub repository at https://github.com/dsfsi/covid19za.git. We use daily incident data arising from the South Africa testing and reporting protocol. South Africa has conducted 6,742,853 COVID-19 tests so far. There are two types of COVID-19 tests that are common in South Africa, i.e. the polymerase chain reaction (PCR) test and the rapid antigen test. Tests are conducted in either private (58% of tests) or public health (42%) facilities. There is ready access to testing.

### 2.2 Model description

We adapted an existing Bayesian SEIR epidemiological model [13,14], using South Africa specific demographics where data is available, to project likely scenarios for a second wave of COVID-19 cases in South Africa. We allow our model to reflect South Africa’s context by assuming initial conditions for state variables and values of parameters for prior distributions that are South Africa specific. Details on parameter values and sources/justification are shown in Table 1. The model reflects the assumption that a fraction of the population is willing and able to observe social distancing measures, such that transmission is considerably reduced among that sub-population and their contacts. The two sub-populations (here called distancing and non-distancing, respectively) are further subdivided into susceptible (*S*), exposed pre-symptomatic (*E*_1_), exposed infectious (*E*_2_), symptomatic and infectious (*I*), quarantined or isolated (*Q*) and recovered or non-transmitting (*R*) individuals. For each of the state variables of the non-distancing population, there is a corresponding state variable with subscript *d*, for the distancing sub-population. The model is a system of first order ordinary differential equations (ODEs) (1).

**Table 1:** Parameter description and values. The fraction of the population engaged in distancing is *e* = *u*_*r*_*/*(*u*_*r*_ + *u*_*d*_).

| Symbol | Definition | Specified/fitted value & Justification |
| --- | --- | --- |
| $N$ | Total population | 57,780,000 (specified) [16] |
| $D$ | Mean duration of the infectious period | 5 days (specified) [17,18] |
| $k_1$ | (time to infectiousness)$^{-1}$ ($E_1$ to $E_2$) | 0.2 days$^{-1}$ (specified) [19,20,21] |
| $k_2$ | (time period of pre-symptomatic transmissibility)$^{-1}$ ($E_2$ to $I$) | 1 days$^{-1}$ (specified) [20,21] |
| $q$ | Quarantine rate | 0.05 (specified) [22] |
| $u_d$ | Rate of people moving to physical distancing | 0.1 (specified) [23] |
| $u_r$ | Rate of people returning from physical distancing | 0.02 (specified) [23] |
| Shape | Weibull parameter in delay-to-reporting distribution | 1.73 (1.60–1.86 95% CI) (specified) [13] |
| Scale | Weibull parameter in delay-to-reporting distribution | 9.85 (9.30–10.46 95% CI) (specified) [13] |
| $f_2$ | Fraction of normal contacts during physical distancing | 0.36 (0.27–0.42 97.5% CI) fitted |
| $\phi$ | Inverse dispersion from negative binomial (NB2) observation model | 6.73 (3.39–12.37 95% CI) fitted |
| $\psi_r$ | Proportion of tested and reported cases on day $r$ | 0.6 specified |
| $R_0$ | Basic reproduction number | Lognormal(log(2.6), 0.2) specified |
| $I_0$ | Fraction of infected individuals in the population at an initial point | Lognormal(log(8), 1) specified |

The following set of ODEs describes the dynamics of the non-distancing sub-population:

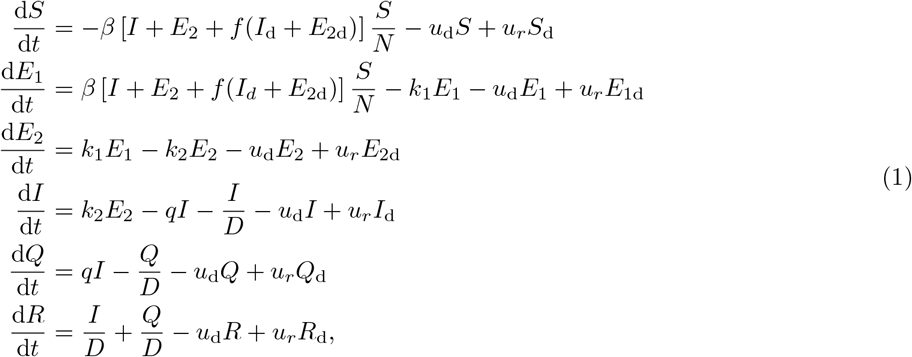

where *β* is the rate of transmission, *D* and *f* are the mean duration of infectiousness and distancing level, respectively. The rates at which individuals move from distancing to non-distancing and vice-versa are *u*_d_ and *u*_*r*_. *k*_1_ is the rate of movement from the exposed pre-symptomatic state to the exposed infectious state, *k*_2_is the rate at which an individual develops symptoms after being infectious but asymptomatic, and *q* is the quarantine or isolation rate of symptomatic individuals. Further parameter descriptions are given in Table 1.

The system of ODEs analogous to (1) for the distancing sub-population is as follows:

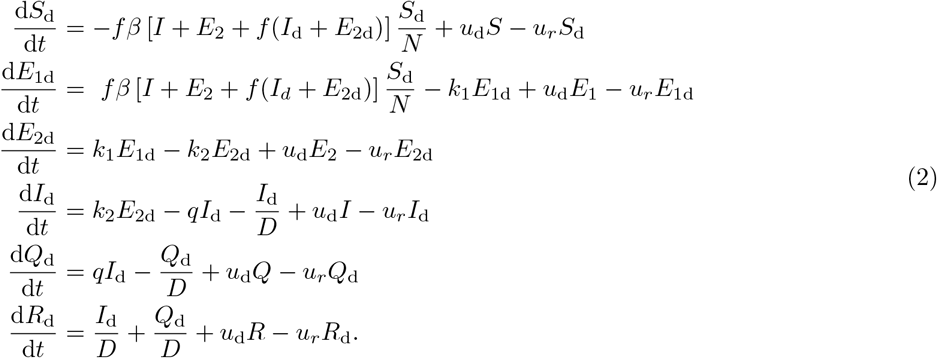

Individuals move between the social distancing and non social distancing sub-populations. The fraction *e* engaging in social distancing can either be estimated from survey data on prevalence of physical distancing, or assumed (a consequence of the rates to and from the distancing compartments) from the model. Here, since there are no data available on the prevalence of physical distancing in South Africa, we assumed a prior beta distribution *β*(0.8, 0.05) for *e*. Full details on parameter values and descriptions are presented in Table 1.

The impact of social distancing measures is assessed by estimating a time varying parameter *f* which measures the fraction of remaining contacts, where the reduction is due to adherence to distancing measures. *f* takes values between 0 and 1; high *f* values indicate low levels of distancing adherence in the population, while low values suggest high compliance with physical distancing measures. The model assumes that there is a background unobserved epidemic that follows the differential equations described above. Furthermore, we assume that only a fraction *ψ*_*r*_ of individuals who develop symptoms are tested and reported daily. Since anyone who has any reason to believe they are positive are encouraged to get tested, and tests can be obtained easily in private health facilities, we believe that the ascertainment rate of symptomatic individuals is at least 60%. We acknowledge that case data arising from testing does not present a full description of the underlying transmission, but if the testing rate is relatively consistent over time, then the reported cases will reflect incidence; furthermore, we do not have data to estimate ascertainment through time.

By incorporating a Weibull distributed delay between onset of symptoms and reporting, and right-censoring (maximum delay of 45 days; see supplementary information in [13]), the model gives a likelihood for the daily number of reported cases given the model and sampling parameters. Priors for the basic reproduction number *R*_0_ (non-distancing; the average number of secondary infections an infected individual is expected to generate during the period of their infectiousness in a wholly susceptible population, largely in the absence of control measures), and the initial fraction of the the population that are infected (*I*_0_), are log-normal with parameters given in Table 1. Our *R*_0_ priors are consistent with early *R*_0_ estimates for COVID-19 in South Africa [15]. The choice of the *I*_0_ prior follows the assumption used for British Columbia in [14], where case numbers were evidently small before the start point of the data. The South Africa data we use starts from the 5 March 2020 when the first case of COVID-19 was confirmed in the country. Our assumption that case numbers are small prior to 5 March is reasonable. Moreover, the parameters for the prior distributions that we use yield a good fit to data. We do not have any data on delay between onset and reporting in South Africa, so we assume that reporting delay is similar to that of British Columbia. Our estimates of *f* and other parameters depend on the assumed priors. An extensive sensitivity analysis in [13] found that conclusions about estimates of the impact of distancing, and the case trajectories, were robust to the assumed ascertainment fraction and other model parameters (though the posterior *R*_0_ was not).

A full description of the model can be found elsewhere [13,14]. The sensitivity of model output to input parameters, including model calibration and validation are presented in [13]. The model is available as an R package *covidseir*, and it can be accessed in the GitHub repository: https://github.com/seananderson/covidseir

## 3 Results

Where available, we use South Africa specific information in our model. Following [13] we estimate the threshold fraction *f* to be 0.469, 95% CI [0.467, 0.471]. Below this the growth rate of the epidemic is negative. At the threshold, the growth rate is 0; above the threshold the growth rate will be positive, and the epidemic will grow exponentially. During the first lockdown we estimate *f* to have been 0.36 95% CI [0.27,0.45], commensurate with declining cases. Figure 1 shows the posteriors of the estimated parameters, with *R*_0_ between 2.5 and 4 and a majority of the population (fraction *e*) participating in distancing (as would be expected in a widespread lockdown). There are some trade-offs. For instance, higher priors for *I*_0_ can lead to lower estimates of *R*_0_. There is also a trade-off between the fraction of the population that are observing social distancing (*e*) and the impact of distancing (*f*), with lower *e* requiring lower *f* to achieve the same prevalence. The reproduction number *R*_0_ is also sensitive to the incubation period and the length of infectiousness period. Lower *R*_0_ and shorter incubation and infectiousness periods will fit the growth rate in a similar way to a higher *R*_0_ and longer incubation and infectiousness periods. The model output depends more on *e* than on the rate at which individuals move between the two sub-populations. However, we find that the fraction *f* is relatively robust to these assumptions, as are the case trajectories under various scenarios. The trade-offs are shown in Figure A.2 in the Appendix. See also [13] for a more detailed discussion.

**Fig. 1:**
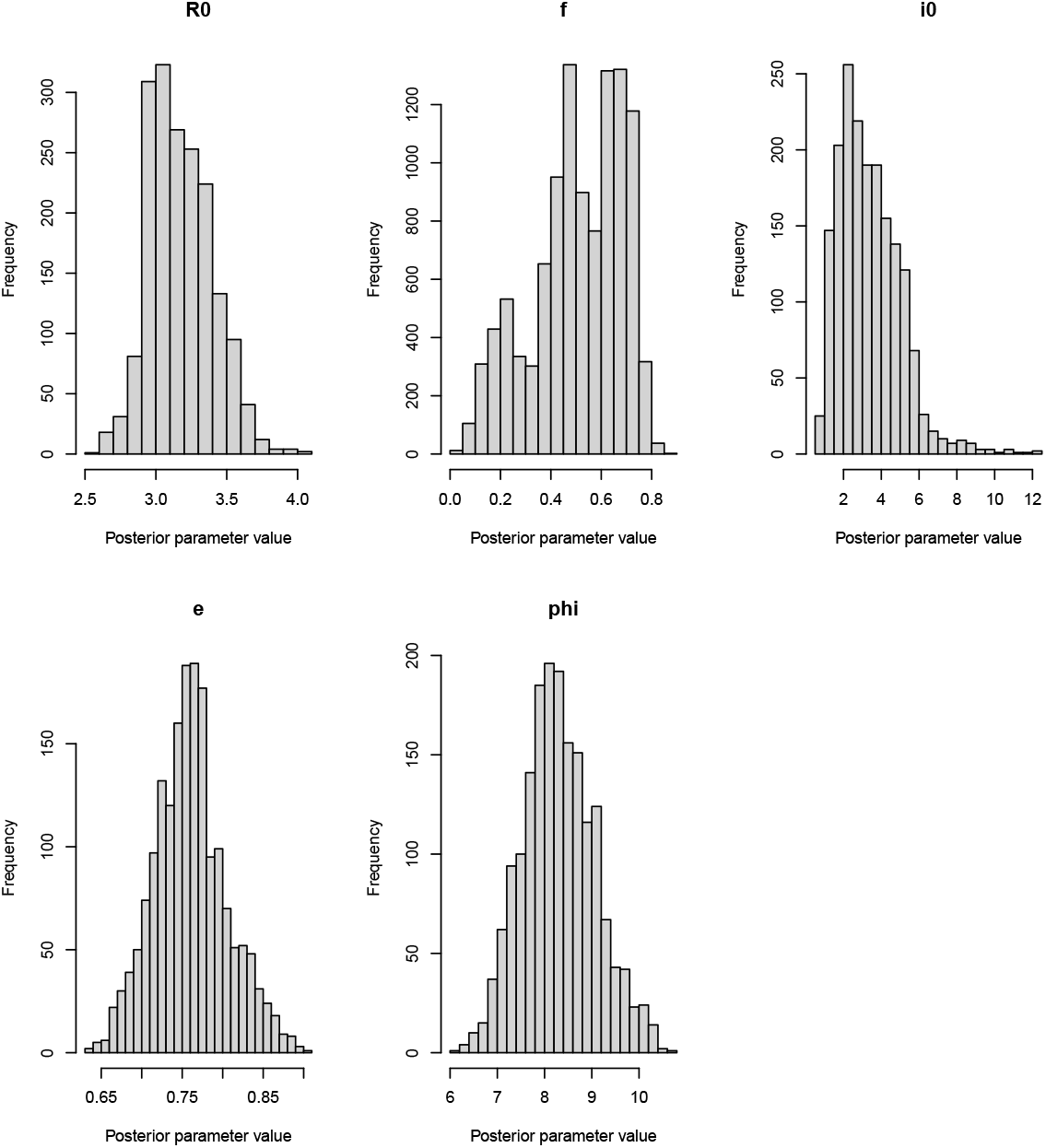
Posteriors of estimated parameters from the model: *R*_0_ is the reproduction number (accounting for quarantine/isolation), *f* is the physical distancing parameter, *I*_0_ is the fraction of the initial population that is infected, *e* is the fraction of the population that is observing physical distancing and phi (*ϕ*) is the dispersion parameter.

### 3.1 Projection of COVID-19 second wave with different levels of contact rate

We project daily reported cases over the next 40 days starting from 29 December 2020, for three levels of contact among those distancing (this reflects the strength of distancing measures and practice). First, the baseline scenario assumes that the current level of contact is sustained over a 40-day period. Secondly, we assume that the current level of contact is reduced by 35%, and thirdly, that the current contact rate is increased by 30% (Fig 2). We note that the latter is unlikely given the current spike in the number of reported cases in the country, and the recently announced adjusted alert level 3 lockdown. Hence, the first and second scenarios are more probable, since contact rates are expected to reduce considerably during the lockdown period. Our model suggests that if the current contact rate is maintained, daily reported cases will continue to rise exponentially with more than 40,000 daily reported cases before the end of January 2021. This would have more than doubled the number of reported cases at the peak of the first wave. The situation will be worse if the current contact rate is increased in form of further relaxation of the current lockdown restrictions. In the other hand, if measures are implemented such that the current contact rates can be reduced to 65% of current rates or less, cases will start to peak after about two weeks from when the lockdown restrictions are implemented, and will continue to decline, provided the reduced contact rate is sustained.

**Fig. 2:**
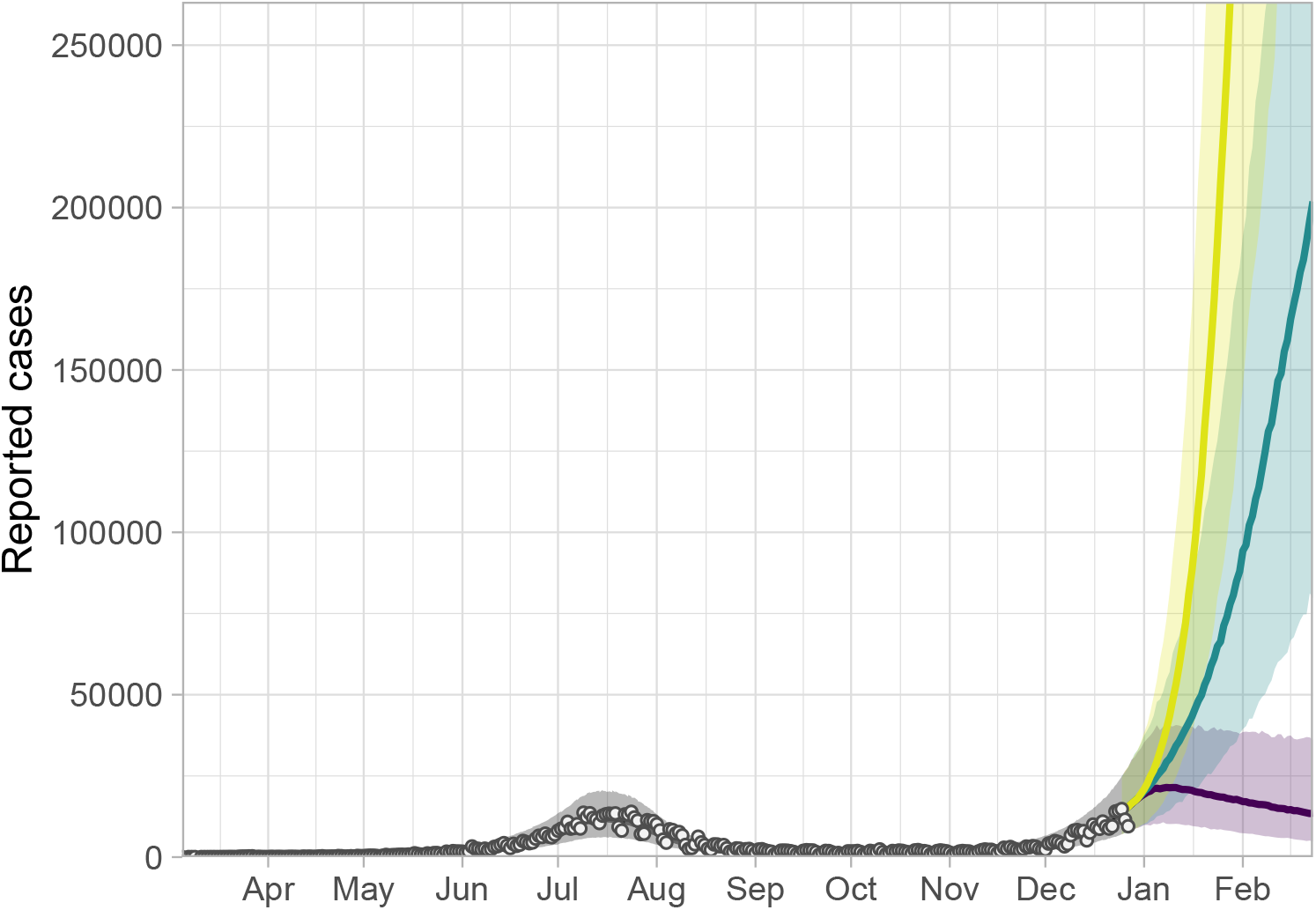
Model fit and projection of the second wave of COVID-19 cases in South Africa for different levels of contacts: dots are the reported case numbers. Solid lines show the model fit and model projections. The yellow, green and purple lines indicate the median of projected case numbers when current rate of contact is increased by 30%, maintained at current levels, or reduced to 65%, respectively. Ribbons represent 90% credible intervals.

These results underscore the need for urgent implementation of serious measures as soon as possible to achieve the net 35% reduction in contact rates. We estimate that the current average contact among those distancing corresponds to an *f* of approximately 0.68 with 95 % CI (0.67, 0.72) of normal contacts. Strict and targeted measures will be needed nation wide to achieve significant reduction in contact to levels that will be sufficient to slow down the growth of the epidemic. A similar conclusion on the need for strict restriction measures to prevent large outbreaks was reached for other African countries [7].

### 3.2 Possible impact of lockdown restrictions on case numbers

The government of South Africa announced adjusted level 3 lockdown restriction on 28 December 2020 and the implementation commenced at midnight on the same day. Restrictions are expected to last until cases are brought under control. We assess the impact of this level 3 lockdown on reported case numbers by reducing *f* values to 0.36 (which is our estimate during level 3 restrictions when cases were declining in the the first wave) (Fig. 3). We find that a 2-week level 3 lockdown restriction will only achieve a temporary reduction of cases starting during the second week of January 2021. After the initial decline, if restrictions were lifted, exponential growth of case numbers would resume on approximately 17 January 2021. By 15 February 2020, daily reported cases could likely reach approximately 50,000. As of 20 December 2020, South Africa was reporting about 10,000 cases per day, and many provinces are already reporting a huge pressure on their hospital capacity, which could be exceeded soon if urgent measures are not taken. With close to 50,000 cases per day by middle of February as predicted by our model, the health care system would likely have been overrun, leading to a serious public health crises.

**Fig. 3:**
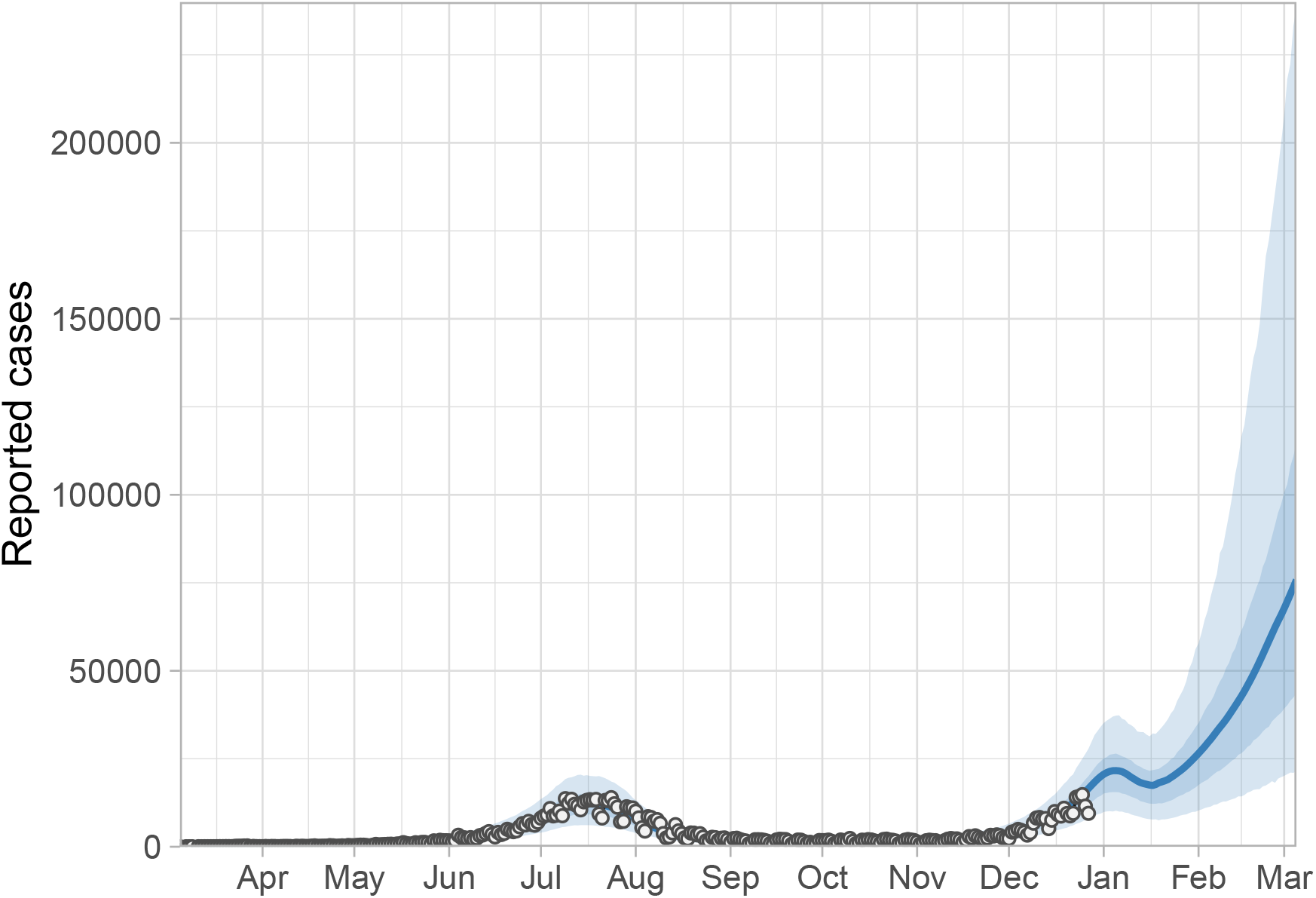
Model fit and projection of second wave of COVID-19 cases in South Africa under 2 weeks level 3 lockdown restrictions. Dots are the reported case data and the solid line is the mean of the projected daily number of cases. Ribbons represent 50% and 90% credible intervals.

Furthermore, we analyse the impact of extended lockdown restrictions, and project how long restrictions will have to last before cases can be brought under control. Our projection predicts that it will take several months of strict lockdown restrictions, if these are the primary means of control, to bring cases down to less than 1,000 per day (Fig. 4).

**Fig. 4:**
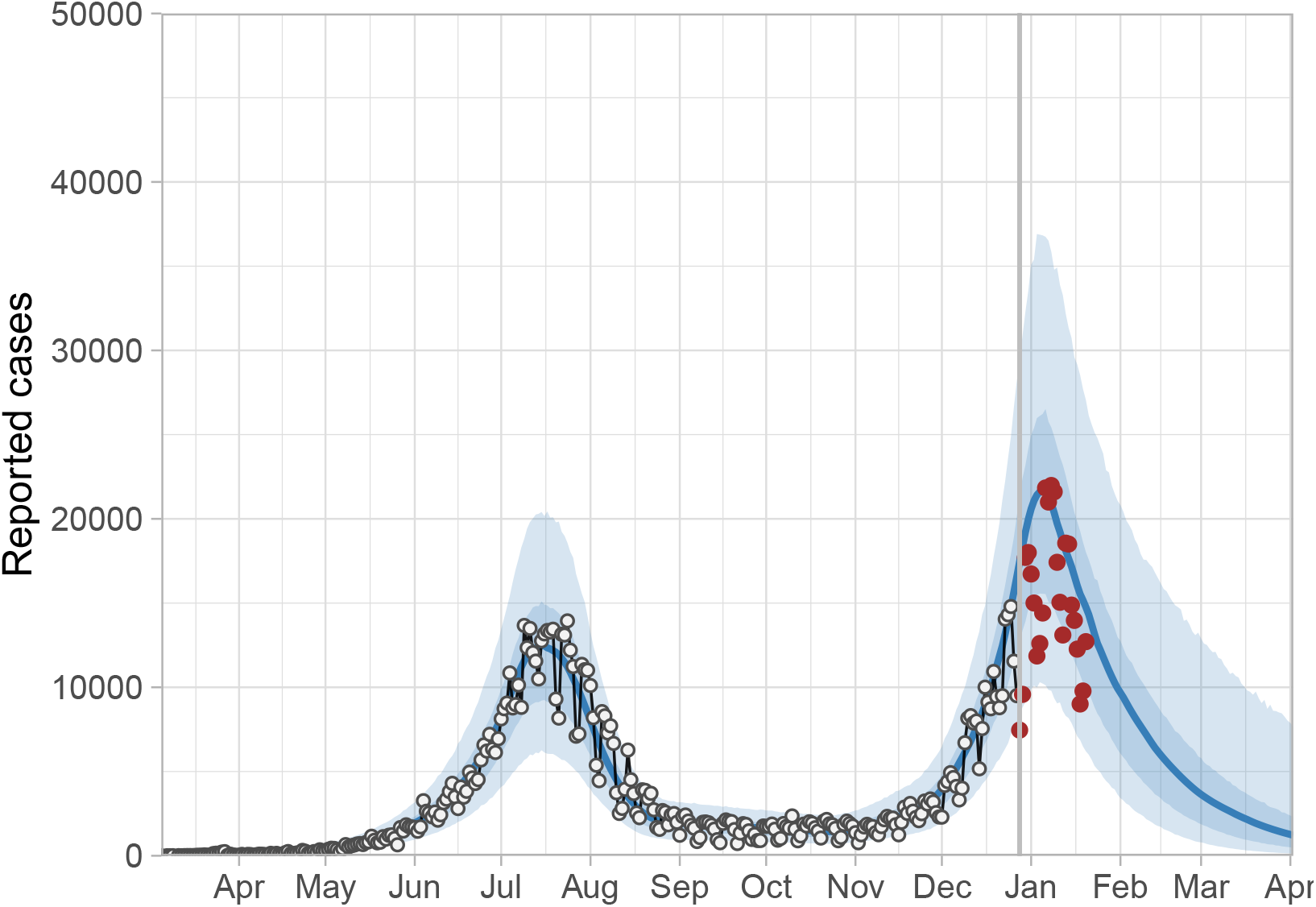
Model fit and projection of the second wave of COVID-19 in South Africa under extended level 3 lockdown restrictions. Dots represent daily reported cases and the solid line is the mean of the projected daily case numbers. Ribbons represent 50% and 90% credible intervals. The grey vertical line indicate the last data point in the data we used for our projection. The dark brown dots are the reported cases after our last model fits.

Strict lockdown restriction will have to be in place from 29 of December to the end of March 2021 before cases can return to the levels observed during the period between the first and the second wave (in the absence of substantial changes to testing, tracing and other COVID-19 control measures). The impacts of such hard restrictions on the economy, inequality and the general well-being of the population are well documented [24,25,26].

## 4 Discussion

Our model results suggests that the current lockdown restrictions, if properly implemented, could help slow down the growth of the epidemic. However, short-term lockdown that lasts for only a few weeks will have only a short-term impact as cases will rise again when restrictions are relaxed in the absence of a carefully planned and properly executed exit strategy [27]. Short-term lockdown restriction should not be viewed as an effective measure with long-term efficacy against COVID-19 transmission. Other studies have drawn similar conclusions, and indeed repeated resurgences following shutdowns and temporary measures have been observed worldwide [28]. For instance, a study that made short-term COVID-19 case predictions for India, Mexico, South Africa and Argentina, predicted that cases will rise in South Africa when restrictions are lifted during the first wave [9].

The success of the lockdown as predicted by our model is predicated on the ability of the lockdown restrictions to reduce contacts to levels close to those that we assumed/predicted in our modelling exercise. We caution that our model should be interpreted with the model assumption around, for example, testing and reporting protocol, in mind. This might change suddenly due to backlog of tests and/or a shift in government testing policy. When these happen, it might influence our model projections. Our model accurately predicted the peak of the second wave, and the reported cases, from the date of our last model fit up until now, fall within the uncertainty bounds of our model projections (Fig. 4). Generally, we are confident that our model projections are reasonable, and robust to a number of modelling assumptions. The sensitivity of the model to all input parameters has been analyzed previously [13]. Additionally, there are uncertainties around parameter values in our model due to lack of detailed high resolution data on, for instance, contact rates during and after the first wave, and/or a detailed line list with information about the course of infection, isolation and reporting.

The second wave of the pandemic in South Africa has already exceeded the peak of the first wave. And we predict that cases will continue to rise until contact rates are reduced to below 65% of current values as of 28 December 2020. The health system could be overwhelmed within a few weeks of reopening. On the other hand, the negative impact of hard prolonged lockdown could be devastating on many fronts. There is an urgent need for multidimensional approaches to the fight against COVID-19 in South Africa. For instance, level 3 lockdown could be sustained and strictly implemented to keep contact rates low until the middle of February 2021, when case numbers would have dropped to about 5000 cases par day. Testing and contact tracing could be concurrently strengthened to ascertain more cases, such that cases and their contacts can isolate or quarantine. Wider testing could support identifying individuals before their period of infectiousness begins, and rapid testing can to allow tests’ turnaround time to fall within 24 hours [29]. This will allow those who test positive to start self-isolation more quickly and drastically reduce onward infections. Extended lockdown restrictions are unpalatable, especially for those who have already been impacted negatively by the first wave. Government may be able to provide palliative support to the most vulnerable for them to be more willing to comply with the lockdown restrictions, and to quarantine or isolate when they are aware that they might have been exposed. These approaches can limit transmission and will hopefully allow gradual decline in case numbers until vaccines are available for roll-out in the country.

A major emerging concern is the discovery of a new variant of SARS-CoV-2 (501Y.V2) that is rapidly spreading across the country. Not much is known currently about this new variant, but preliminary investigations suggest that it could be more transmissible because of its association with higher viral loads [30], although higher viral loads can also be observed simply because in a growing epidemic, most individuals observed were infected recently [31]. Another recent study suggests that the 501Y.V2 variant shows changes in severity, and is either more transmissible and/or is able to escape previously acquired immunity [32]. It is still unclear if the new variant will be associated with more severe disease or will lead to more fatal outcomes compared to the previous variants that dominated during the first wave [30]. As more data emerges it will be possible to compare the variants’ reproduction number, virulence and transmissibility to baseline COVID-19 values, and to explore the implication of the new variant for vaccination, testing, therapeutics and other epidemiological implications.

With the discovery and regulatory approval of several effective vaccines (e.g. Pfizer-BioNTech, Moderna, Sinopharm, and Oxford-AstraZeneca), procuring and distributing vaccines should be of high priority. The government of South Africa has announced that vaccines will be available for use against COVID-19 in the second quarter of 2021. Until effective vaccines are available and accessible in the country, lockdown restrictions will only provide a temporary measure against COVID-19 in South Africa. Extended hard lockdown restrictions are not tolerable and are too expensive to be used as a standalone measure against COVID-19 transmission. Extended hard lockdowns will have a very high cost both in economic terms [26] and in the impact on health and broader society [24]. Given that reducing transmission through vaccination is many months away, such lockdowns may be best used as a tool to slow the spread of COVID-19 while strengthening the health care system, increasing efforts towards procurement and deployment of vaccines in conjunction with other measures such as mass rapid testing, and ensuring compliance with ongoing physical distancing measures and mask mandate.

## Data Availability

We use public data available at https://github.com/dsfsi/covid19za.git

## Appendix

The appendix contains the Markov Chain Monte Carlo (MCMC) trace plot and the pairs plot, showing the tradeoffs involved in our assumptions about priors for our model parameters.

**Fig. A.1:**
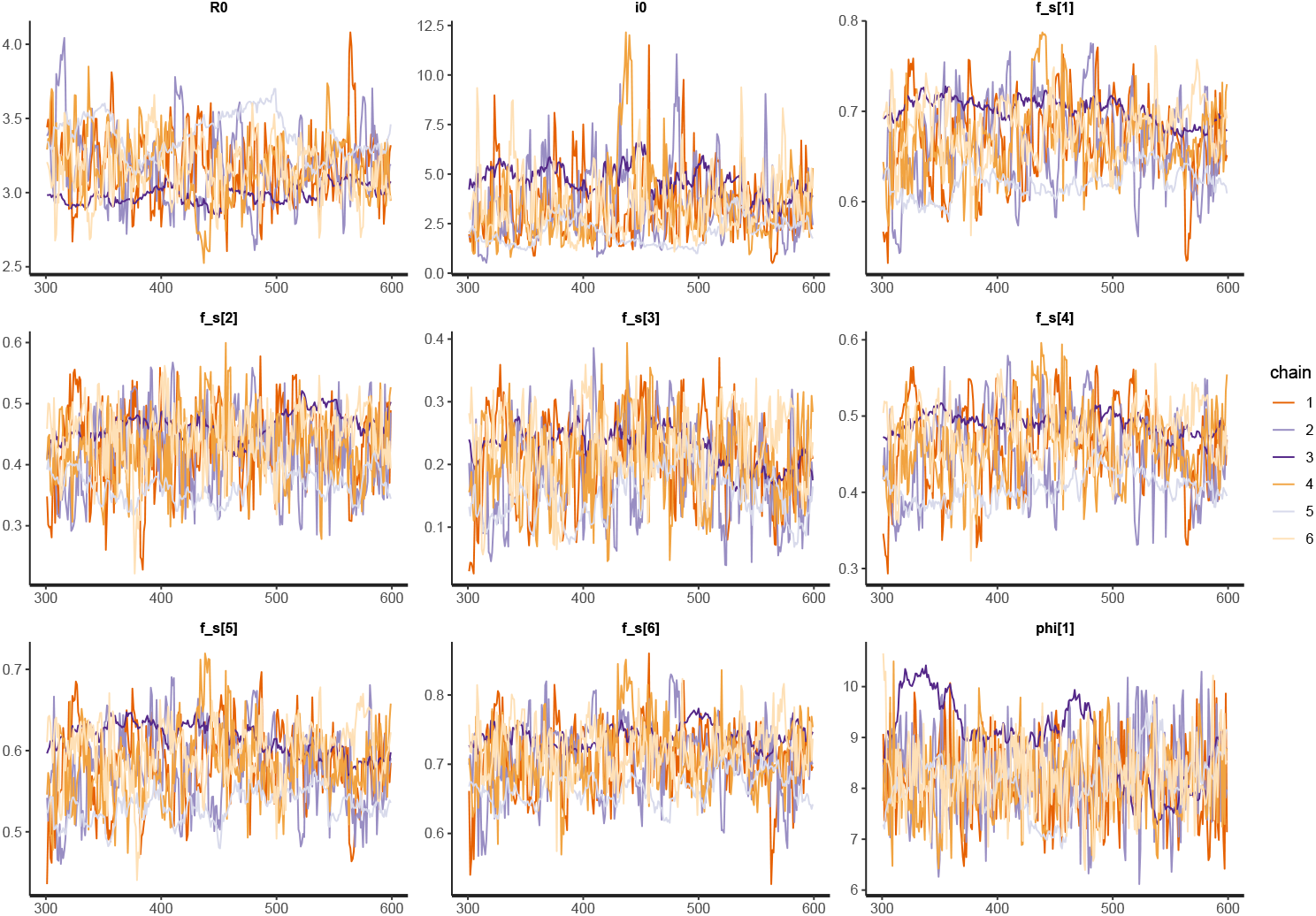
Trace plots of Markov Chain Monte Carlo (MCMC) samples from parameter distribu- tions to test for chain convergence

**Fig. A.2:**
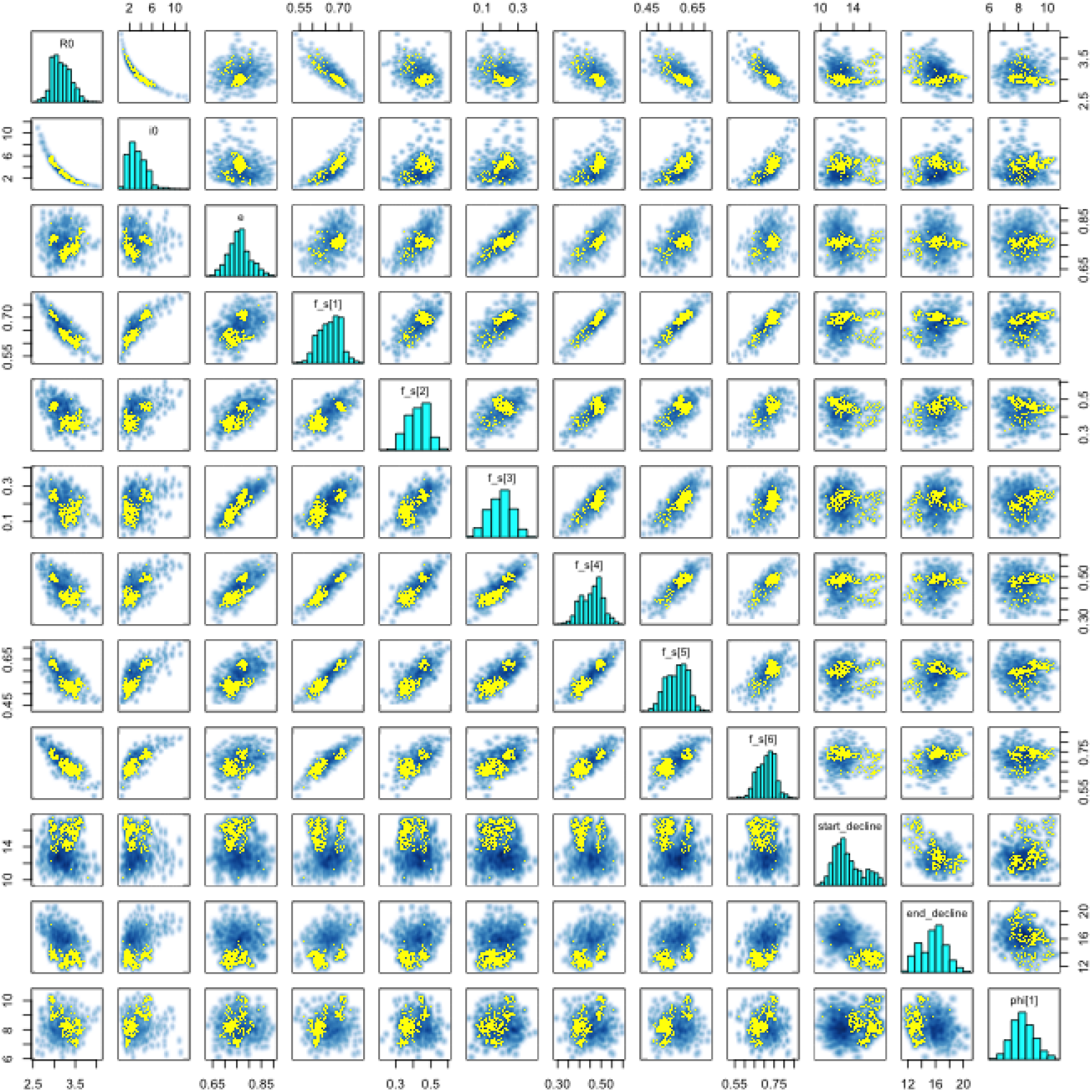
Pairs plot of the Markov Chain Monte Carlo (MCMC) samples for the estimated parameters.

